# Proteomics signature of physical activity and risk of multimorbidity of cancer and cardiometabolic diseases

**DOI:** 10.1101/2025.03.26.25324689

**Authors:** Michael J. Stein, Hansjörg Baurecht, Patricia Bohmann, Reynalda Cordova, Pietro Ferrari, Béatrice Fervers, Christine M. Friedenreich, Marc J. Gunter, Laia Peruchet-Noray, Diana Wu, Michael F. Leitzmann, Vivian Viallon, Heinz Freisling

## Abstract

**Background:** Cancer, cardiovascular diseases (CVD), and type 2 diabetes (T2D) may co-occur, a condition referred to as multimorbidity. Physical activity is inversely associated with each of these diseases; however, the biologic pathways underlying these relationships remain incompletely understood.

**Methods:** In 33,806 UK Biobank participants, we derived a proteomic signature (high-throughput panel of 2,911 proteins assessed by Olink array) of moderate-to-vigorous physical activity using linear and LASSO regressions in a two-step procedure to prospectively assess associations with physical activity-related cancers (1,108 cases), CVD (3,445 cases), T2D (1,363 cases), as well as progression to multimorbidity (420 cases). Multivariable Cox regression estimated hazard ratios (HRs) and 95% confidence intervals (CIs) for each identified protein, as well as their linear combination (proteomics signature score), separately for each outcome and with adjustment for physical activity. Pathway enrichment analysis and protein-protein interaction networks were used to gain insights into the systemic interplay of the identified proteins.

**Results:** After correction for multiple testing, 223 proteins were selected in the physical activity signature. Proteins involved in food intake, metabolism, and cell growth regulation (e.g., LEP, MSTN, TGFBR2) were inversely associated with physical activity. Proteins involved in immune cell adhesion and migration, as well as cartilage and muscle integrity (e.g., integrins, COMP, MYOM3) were positively associated with physical activity. Several proteins upregulated by physical activity were inversely associated with disease risk (e.g., integrins, PI3, CLEC4A for cancer risk, or LPL, IGFBP1, LEP for T2D risk). Similarly, various proteins were downregulated by physical activity and positively associated with disease risk (e.g., CD38, TGFA for CVD risk). For multimorbidity, proteins inversely related to physical activity generally aligned with expected risk patterns, while positively associated proteins exhibited mixed effects, with inverse and positive associations. The proteomics signature score was inversely associated with the risk of cancer (HR per interquartile range: 0.87; 95% CI: 0.78, 0.96) and T2D (HR: 0.66; 95% CI: 0.60, 0.72), after adjustment for physical activity, but not with CVD (HR: 0.93; 95% CI: 0.85, 1.03) and progression towards multimorbidity.

**Conclusions:** These findings suggest that the inverse relationships between physical activity and risk of major chronic diseases may be explained by the maintenance of tissue integrity and the proper regulation of immune and metabolic processes. Further studies are needed to determine the causal nature of these associations.

## 1. Background

Cancer, cardiovascular diseases (CVD), and type 2 diabetes (T2D) are the leading causes of death globally.^1^ Moreover, the cancer burden is projected to increase by 47% by 2040.^2^ Similarly, global cases of CVD and T2D nearly doubled between 1990 and 2019.^3,4^ Alarmingly, four out of ten individuals with cancer have at least one other chronic disease, most commonly CVD or T2D.^5^ This co-occurrence of multiple conditions is referred to as multimorbidity^6^ and may be explained by shared risk factors across diseases, such as low physical activity.^7^ In fact, physical activity is related to decreased risk of cancer, CVD, and T2D^8^ as well as multimorbidity.^9^

The biological pathways through which physical activity may reduce the risk of any of these diseases are hypothesized to involve improvements in insulin sensitivity, reductions in chronic low-grade inflammation, and modulation of sex hormone activity.^10^ However, the exact biological pathways remain incompletely defined and poorly understood.

Contemporary technological advancements in proteomics have facilitated high-throughput proteomic profiling in large cohorts, enabling the analysis of thousands of circulating proteins and their associations with lifestyle and health. It is well-established that acute physical activity affects several hundred circulating proteins.^11,12^ In addition, the proteome is relatively stable over time (e.g., compared to more transient metabolic states) and is directly linked to biological pathways and physiological processes.^13^ Consequently, long-term physical activity may be reflected in blood protein concentrations, making it a useful tool for epidemiologic studies of the biochemical mechanisms linking physical activity and health.

Three previous cohort studies have analyzed physical activity in relation to the proteome. One smaller study (n=897) identified five proteins that were associated with physical activity,^14^ while a larger proof-of-concept study (n=11,695) found 65 proteins linked to physical activity.^15^ In 2025, a UK Biobank study (n=39,160) identified 41 proteins consistently associated with various physical activity measures, including self-reported and device-based physical activity, and linked these proteins to dementia risk.^16^ These studies have demonstrated the potential to uncover biochemical processes related to physical activity, some of which may influence health. However, no study has yet comprehensively examined the link between physical activity-related proteins and cancer or cardiometabolic health.

In this study, we assessed large-scale proteomic data to derive a proteomics signature of physical activity and investigate its association with cancer, CVD, T2D, and multimorbidity.

## 2. Methods

### Study design and participants

The UK Biobank is a prospective cohort study that enrolled 502,134 UK participants aged 40 to 69 years between 2006 and 2010. The study collected sociodemographic, lifestyle, and extensive phenotypic data using touchscreen questionnaires, interviews, physical and functional measurements, and biomaterial collection. Ethical approval was obtained from the North West Multi-Centre Research Ethics Committee, and all participants provided written informed consent.^17^ The UK Biobank Pharma Proteomics Project (UKB-PPP) is a precompetitive consortium that generated protein measurements using the Olink Proximity Extension Assay, capturing 2,923 unique proteins in 53,026 participants. Details of the inclusion process for the UKB-PPP are described elsewhere.^18^ In brief, an initial sample of 6,229 participants were pre-selected by UKB-PPP members, and the remaining representative samples were selected from the UK Biobank, stratified by age, sex, and recruitment center.

After exclusion of prevalent cases of cancer, CVD, and T2D as well as incident cases during the first year of follow-up (n=8,743), participants with missing exposure or covariate data for protein identification (n=8,409), and those with a substantial proportion of missing protein data (>50% missingness, n=2,055), a total of 33,806 participants were included in the proteome– disease analyses (see Supplementary Figure 1 for a flowchart of participant exclusions).

### Physical activity measurement

Physical activity was assessed using the International Physical Activity Questionnaire (IPAQ) Short Form,^19^ which captures weekly frequency and daily duration (minimum 10 minutes) of walking, moderate, and vigorous physical activity over the previous four weeks. Following the IPAQ evaluation protocol,^19^ Metabolic Equivalent of Task (MET) values from the Compendium of Physical Activities developed by Ainsworth and colleagues^20^ for moderate (4.0 METs) and vigorous (8.0 METs) activities were multiplied by the respective frequency and duration to estimate total MET-hours per week (MET-hours/week) of moderate-to-vigorous physical activity. No distinction was made between recreational and occupational activities. We focused on moderate-to-vigorous physical activity due to its purportedly stronger physiological effects compared to activities of light intensity.

### Protein measurements and proteomic data processing

The Olink assay technology and the analyses of the UKB-PPP are detailed elsewhere.^18,21^ In brief, the relative abundance of 2,923 proteins was quantified using antibodies distributed across four 384-plex panels: inflammation, oncology, cardiometabolic, and neurology. Blood samples were assayed in four 384-well plates consisting of four abundance blocks for each of the four panels per 96 samples, using the Olink Explore platform. This platform is based on proximity extension assays, which are highly sensitive and reproducible, with low cross-reactivity. Relative concentrations of the 2,923 unique proteins were measured by next-generation sequencing and expressed as normalised protein expression (NPX) values on a log-base-2 scale. Following a previously published approach,^22^ protein values below the limit of detection (LOD) were replaced with the LOD divided by the square root of two.^23^ Proteins with more than 20% missingness (n=12) were excluded. Missing protein expression levels were imputed using k-nearest neighbors (*k*=10), allowing for up to 50% missingness across proteins and 20% per protein. Finally, each protein was rescaled to have a mean of 0 and a standard deviation (SD) of 1.

### Cohort follow-up and outcome ascertainment

Participants’ vital status was obtained through linkage with health care data and national death registries^24^. Follow-up began at baseline and ended at the date of complete follow-up (December 2016 for Scotland, December 2020 for England, November 2021 for Scotland)^25^, loss to follow-up, or death, whichever came first. For cancer incidence, we focused on the first primary malignant cancers with convincing evidence for an association with physical activity,^26^ including cancers of the breast, colorectum, corpus uteri, bladder, kidney (renal cell carcinoma), esophagus (adenocarcinoma), and stomach (cardia), which were combined into a composite “cancer” outcome. The composite CVD outcome included diagnoses of angina pectoris, myocardial infarction, ischaemic heart diseases, arterial fibrillation, cardiac arrhythmias, heart failure, cerebrovascular diseases and stroke, atherosclerosis, and other peripheral vascular diseases. T2D was identified using the International Classification of Diseases (ICD)-10 code E11. A complete list of all ICD codes for all outcomes is provided in Supplementary Table 1.

Multimorbidity was defined as the presence of any combination of two of the following outcomes: cancer, CVD, or T2D. When two diseases occurred on the same day (n=195), they were separated by one day based on an arbitrary temporal order (T2D, cancer, CVD). To address reverse causation and detection bias, we excluded the first year of follow-up after diagnoses of primary outcomes (exclusion of 394 cases).

### Proteomic analyses of physical activity

We identified proteins associated with physical activity using a two-step procedure. In the first step, we applied separate linear regression models for each protein to screen for physical activity-associated proteins. We used standardized weekly moderate-to-vigorous physical activity as the outcome variable, and all 2,911 protein expression variables were included as predictors, alongside major potential confounders, including sex, age, study center, body mass index, diet, alcohol, and smoking. We assessed potential non-linearity using restricted cubic splines with four knots (0.05, 0.35, 0.65, 0.95 quantiles) and log likelihood ratio tests. In the second step, we applied least absolute shrinkage and selection operator (LASSO) regression to these pre-identified proteins to select the most predictive proteins. Covariates were not penalized in the LASSO model. The entire feature selection process was designed to minimize potential collider bias, which could otherwise distort protein–physical activity associations. We performed LASSO regression with fivefold nested cross validation (five outer fold, five inner folds) for the assessment of the association with physical activity, tuning the regularization parameter lambda across 100 values from the maximum down to 0.1% of that value. Model performance was evaluated using the mean squared error to identify the optimal lambda.

We retained proteins with non-zero coefficients as physical activity-related proteins and created a physical activity proteomics signature for each participant, calculated as the weighted sum of protein levels using the respective ß-coefficients from the LASSO model as weights.

### Protein-protein interaction networks and pathway enrichment analyses

To gain insights into the underlying biologic mechanisms of the identified proteins, we constructed protein-protein interaction networks using STRING^27^ and visualized them with Cytoscape.^28^ To identify proteins with the highest number of interactions, the maximal clique centrality (MCC) method was utilized.^29^

We conducted pathway enrichment analyses to investigate whether physical activity-associated proteins clustered within distinct biological pathways. Using the Gene Ontology resource^30^, Kyoto Encyclopedia of Genes and Genomes (KEGG),^31^ Reactome,^32^ and WikiPathways,^33^ we examined enrichment across biological functions, molecular pathways, and cellular components. Enrichment tests were performed against a background gene set corresponding to all proteins included in the analysis.

### Statistical analyses

We used Cox proportional hazard regression with age as the underlying time metric^34^ to estimate hazard ratios (HRs) and corresponding 95% confidence intervals (CIs) for the associations between individual physical activity-associated proteins, and separately the proteomic signature score, with cancer, CVD, T2D, and multimorbidity. The Cox models were stratified by age at baseline (5-year increments), sex, and country (England, Scotland, Wales), and adjusted for education level (highest, intermediate, lowest, none of those), socio-economic status (Townsend index, categorized using tertiles, missing values coded as missing), smoking (never, former, current), alcohol use (never, former, current), sedentary behavior (0-3h, 4-5h, 6-7h, >8h of daily TV watching, PC use during leisure, and driving), and screening for breast and/or bowel cancer (binary) as categorical variables, as well as physical activity (MET-hours), body mass index (kg/m^2^), and diet (healthy diet score, 0-7 scale)^35^ as continuous variables. Non-linearity was addressed using restricted cubic splines with four knots at the 0.05, 0.35, 0.65, and 0.95 quantiles. Departures from linearity were assessed using log likelihood ratio tests.

In additional analyses, we evaluated whether excluding physical activity adjustment influenced the association between the proteomic signature and disease risk by assessing model fit using likelihood ratio tests, Akaike Information Criterion (AIC), and the concordance index (Harrell’s C-index). Additionally, we computed HRs and 95% CIs for both variables under single and mutual adjustment scenarios to explore potential mechanistic insights.

All P-values to assess exposure outcome associations and for enrichment analyses were adjusted for multiple comparisons using the Benjamini-Hochberg procedure (false discovery rate q-values) with a significance threshold of 5%.

All data processing and statistical analyses were performed using R 4.4.^36^ Cox regression analyses were performed using the *rms* package.^37^

## 3. Results

We assessed proteome–disease associations in 33,806 participants (53% women, median age 57 years [interquartile range: 49 to 63]) (Table 1). During a median follow-up of 11.8 years (interquartile range: 11.1 to 12.5), 1,108 participants developed cancer, 3,445 developed CVD, 1,363 developed T2D, and 814 developed multimorbidity (Figure 1A).

**Table 1.** Baseline population characteristics of the UK Biobank proteomics cohort.

|  |  |  |  |
| --- | --- | --- | --- |
| Characteristic | Overall | Women | Men |
| **N** | 33,806 | 17,898 | 15,908 |
| **Physical activity (MET-hours)** | 27.0 (33.5) | 24.8 (29.9) | 29.4 (37.1) |
| **Age (years)** | 55.8 (8.2) | 55.7 (8.1) | 56.0 (8.3) |
| **Country** |  |  |  |
| England | 29,976 (88.7%) | 15,863 (88.6%) | 14,113 (88.7%) |
| Scotland | 2,452 (7.3%) | 1,312 (7.3%) | 1,140 (7.2%) |
| Wales | 1,378 (4.1%) | 723 (4.0%) | 655 (4.1%) |
| **Body mass index (kg/m<sup>2</sup>)** | 27.2 (4.6) | 26.8 (5.0) | 27.5 (4.0) |
| **Townsend Index** | -1.4 (3.1) | -1.4 (3.0) | -1.4 (3.1) |
| Missing | 42 (0.1%) | 15 (0.1%) | 27 (0.2%) |
| **Education** |  |  |  |
| Highest | 12,394 (36.7%) | 6,289 (35.1%) | 6,105 (38.4%) |
| Intermediate | 7,904 (23.4%) | 4,083 (22.8%) | 3,821 (24.0%) |
| Lowest | 8,812 (26.1%) | 5,052 (28.2%) | 3,760 (23.6%) |
| Other | 4,462 (13.2%) | 2,357 (13.2%) | 2,105 (13.2%) |
| Missing | 234 (0.7%) | 117 (0.7%) | 117 (0.7%) |
| **Smoking status** |  |  |  |
| Never | 19,030 (56.3%) | 10,873 (60.7%) | 8,157 (51.3%) |
| Former | 11,397 (33.7%) | 5,528 (30.9%) | 5,869 (36.9%) |
| Current | 3,379 (10.0%) | 1,497 (8.4%) | 1,882 (11.8%) |
| **Alcohol use** |  |  |  |
| Never | 1,356 (4.0%) | 916 (5.1%) | 440 (2.8%) |
| Former | 1,125 (3.3%) | 626 (3.5%) | 499 (3.1%) |
| Current | 31,325 (92.7%) | 16,356 (91.4%) | 14,969 (94.1%) |
| **Healthy diet score** | 3.6 (1.4) | 3.9 (1.2) | 3.3 (1.4) |
| **Sedentary behavior (hours)** | 4.5 (2.6) | 4.0 (2.3) | 5.0 (2.7) |
| Missing | 4 | 2 | 2 |
| **Cancer screening** |  |  |  |
| Never | 14,714 (43.5%) | 3,608 (20.2%) | 11,106 (69.8%) |
| Ever | 19,092 (56.5%) | 14,290 (79.8%) | 4,802 (30.2%) |
| MET: metabolic equivalent of task |  |  |  |
| Numbers show mean (standard deviations) or N (percentage). |  |  |  |

**Figure 1.**
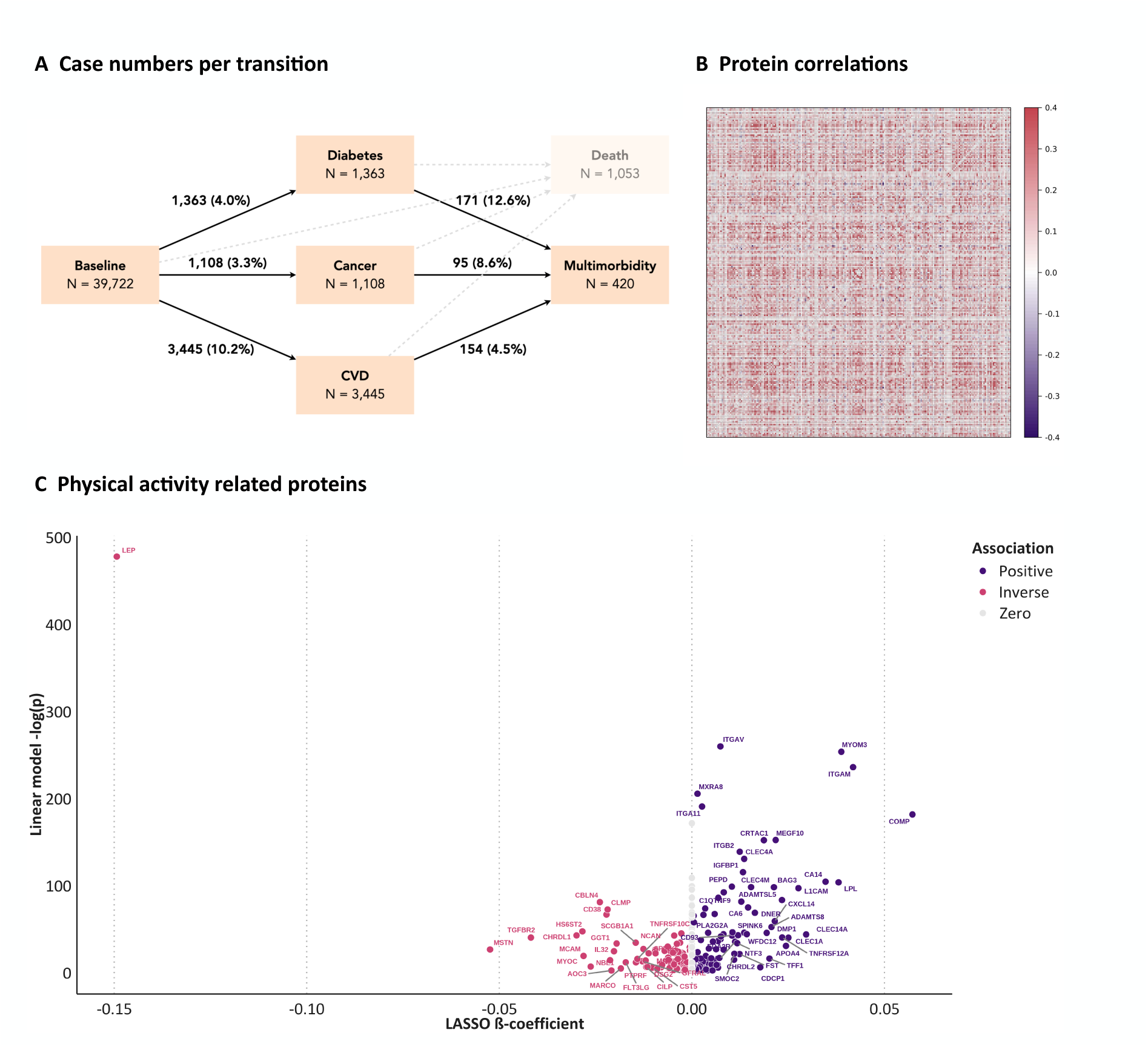
A: Case numbers for each transition from baseline to outcome, from outcomes to multimorbidity as well as death (for completeness). B: Pairwise correlations for each physical activity-related protein. C: Comparison of LASSO ß-coefficients (on the x-axis) and the negative logarithm of the FDR-corrected P-value from linear regression models (on the y-axis) for the 223 identified proteins.

### Physical activity related proteins

In a pre-filtering step using linear regression analyses, we identified 1,000 proteins associated with physical activity. Subsequently, we fitted this subset of proteins with LASSO regression and identified 223 proteins with non-zero coefficients, of which 119 were inversely and 104 positively associated with physical activity (Supplementary Table 2). Pairwise correlations revealed weak to moderate correlations among these proteins (Figure 1B). Among the most strongly inversely associated proteins with physical activity were LEP, MSTN, and TGFBR2, whereas COMP, ITGAM, and MYOM3 showed the highest positive regression coefficients (Figure 1C).

### Protein-protein interaction networks and pathway enrichment analysis

Among the proteins that were inversely associated with physical activity, we identified 69 proteins forming 95 protein-protein interactions, with LEP, IGFBP3, and PTH emerging as the three hub proteins (Figure 2). Among the proteins positively associated with physical activity, 76 proteins were involved in 186 protein-protein interactions. A strong interconnected network of integrin proteins was observed, including the three hub proteins ITGAM, ITGAV, and ITGB2.

**Figure 2.**
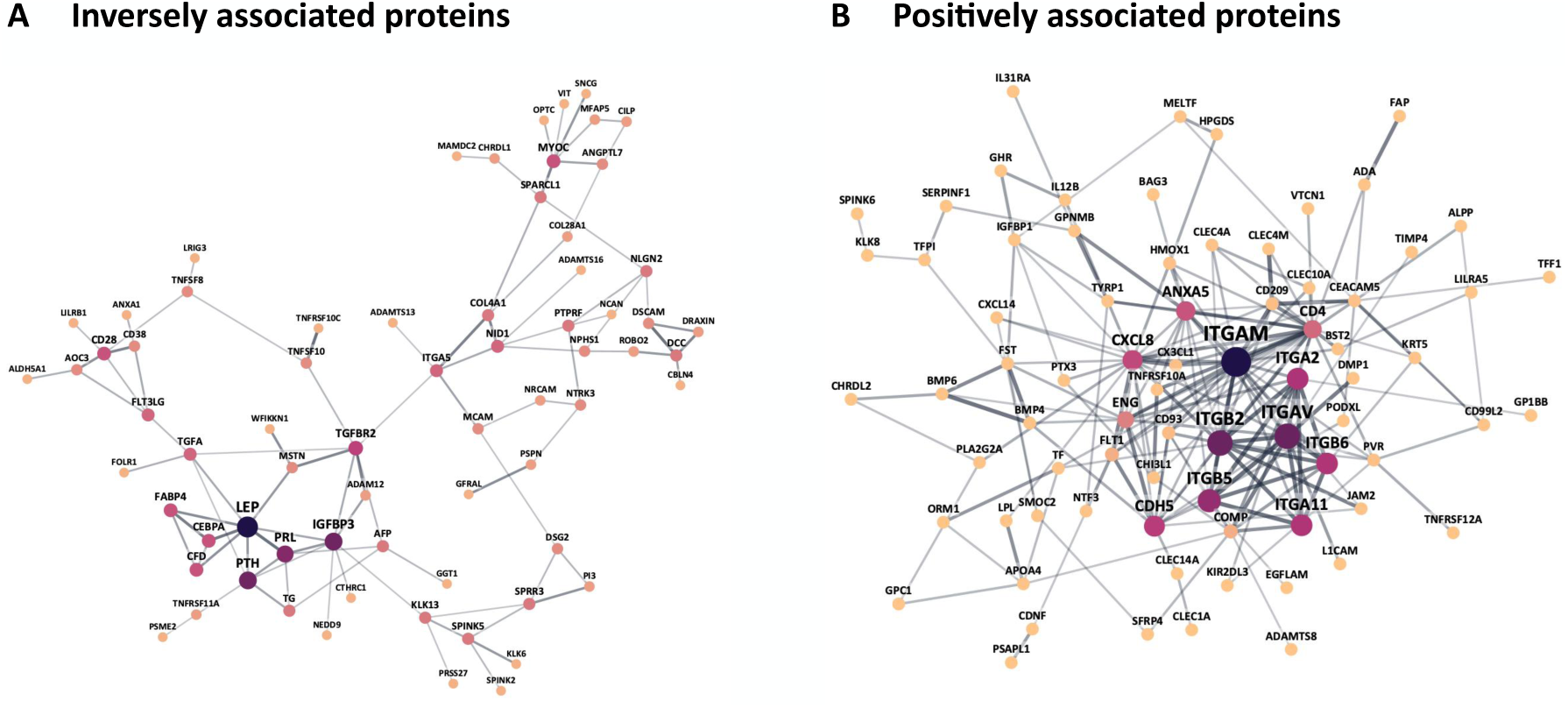
Protein-protein interaction networks for (A) inversely with physical activity related proteins and (B) positively associated proteins. The size of the node reflects the modularity level (MCC method) and the width of the edges reflect the interaction score.

Pathway enrichment analyses revealed that proteins inversely associated with physical activity were enriched in extracellular region components and multicellular organism processes. Proteins positively associated with physical activity were enriched in cell surface and membrane-associated complexes, particularly integrin complexes and receptor signaling structures, suggesting roles in cell adhesion and communication. Molecular functions included integrin and opsonin binding, indicating involvement in extracellular matrix interactions and immune recognition (Table 2).

**Table 2.** Pathway enrichment analyses for inversely and positively with physical activity related proteins.

|  |  |  |
| --- | --- | --- |
| **Category** | **Description** | **q-value** |
| **Inversely associated proteins** |  |  |
| GO Cellular Component | Extracellular region | 0.0017 |
| GO Biological Process | Multicellular organismal process | 0.0181 |
| **Positively associated proteins** |  |  |
| GO Cellular Component | Cell surface | 0.0000 |
|  | Integrin complex | 0.0018 |
|  | Integral component of membrane | 0.0022 |
|  | Intrinsic component of membrane | 0.0022 |
|  | External side of plasma membrane | 0.0041 |
|  | Protein complex involved in cell adhesion | 0.0042 |
|  | Cell periphery | 0.0124 |
|  | Plasma membrane signaling receptor complex | 0.0124 |
| GO Molecular Function | Integrin binding | 0.0057 |
|  | Opsonin binding | 0.0209 |
|  | Protein-containing complex binding | 0.0453 |
| GO: Gene ontology. |  |  |

### Proteome–disease associations

We identified 11 proteins associated with cancer risk after adjusting for risk factors, four of which were inversely and seven positively related. Among the top hits, CHRDL2 (HR per interquartile range: 1.18; 95% CI: 1.10, 1.27, q=8.24×10^−5^) and PI3 (HR: 1.14; 95% CI: 1.06, 1.22, q=5.84×10^−3^) were strongly positively associated with cancer risk, while CLEC4A showed a non-linear inverse association (P-nonlinear=0.007). The proteomics signature was linearly and inversely associated with cancer risk (HR: 0.87; 95% CI: 0.78, 0.96, q=3.31×10^−2^).

For CVD risk, 50 proteins were associated with decreased risk and 48 with increased risk, with EDA2R emerging as the most prominent (HR: 1.21; 95% CI: 1.09, 1.34; q=5.74×10^−24^; P-nonlinear=1.57×10^−2^). The proteomics signature was non-linearly inversely associated with CVD risk (HR: 0.93; 95% CI: 0.85, 1.03; q=8.45×10^−2^; P-nonlinear=0.014); not passing the false discovery rate threshold.

Regarding T2D risk, 63 proteins were inversely associated and 91 were positively associated, with MXRA8 emerging as the top hit (HR: 0.51, 95% CI: 0.48, 0.55; q=1.32×10^−67^). The proteomics signature was linearly and inversely associated with T2D (HR: 0.66; 95% CI: 0.60, 0.72; q=2.23×10^−18^).

Figure 3 shows the HRs for all proteins and the signatures for each transition. Figure 4 additionally displays proteins that were statistically significantly associated (after FDR correction) with at least one outcome. HR curves for linear and non-linear trends are visualized in Supplementary Figure 2 for both the proteomic signatures and the top hits for each transition.

**Figure 3.**
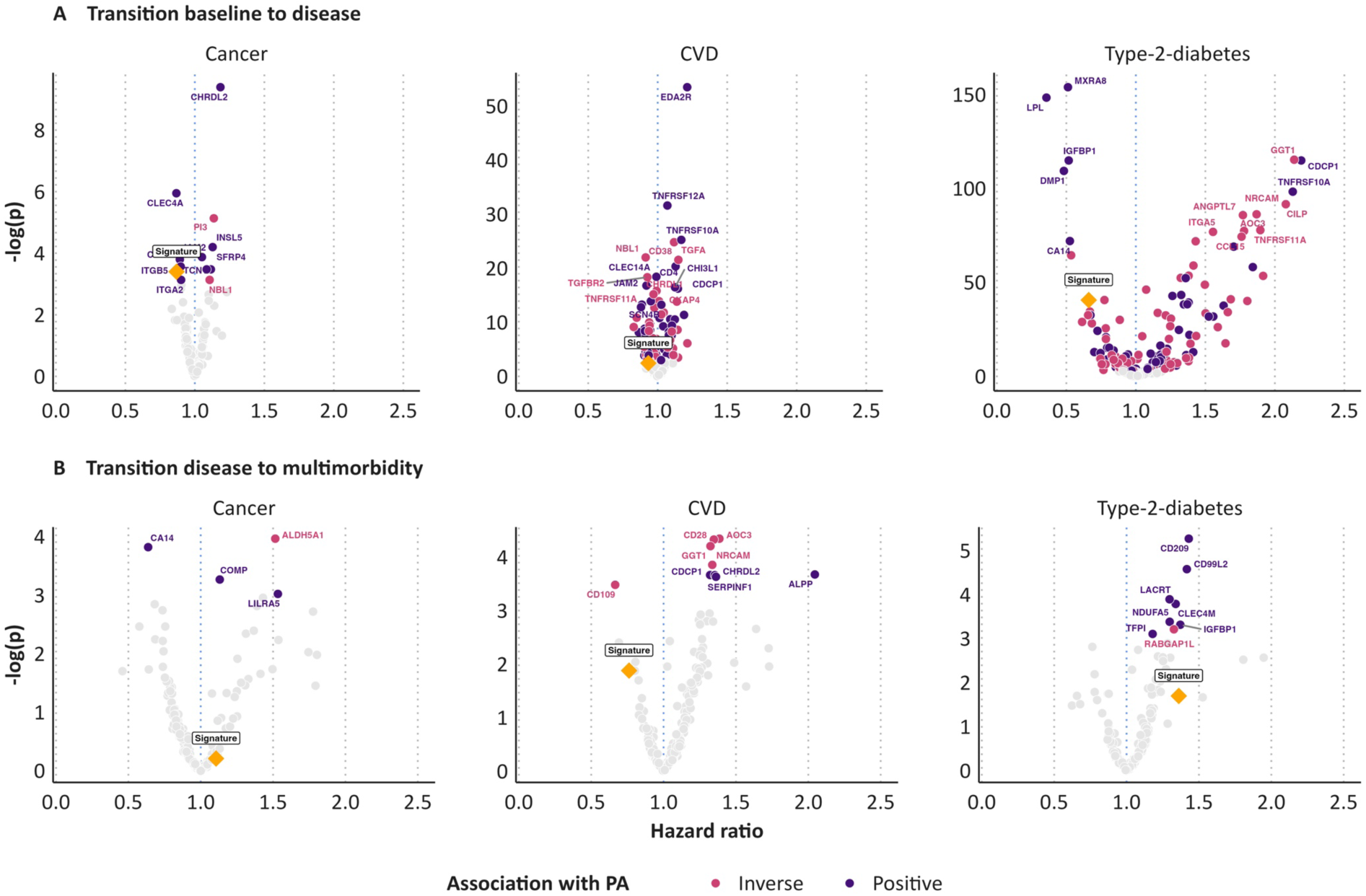
Hazard ratios for all proteins and the proteomics signature for each of the six transitions. Y-axis shows the negative logarithm of the P value after false discovery rate correction. All models were stratified by age at baseline (5-year increments), sex, and country (England, Scotland, Wales), and adjusted for education level (highest, intermediate, lowest, none of those), socio-economic status (Townsend index, categorized using tertiles, missing values coded as missing), smoking (never, former, current), alcohol use (never, former, current), sedentary behavior (0-3h, 4-5h, 6-7h, >8h of daily TV watching, PC use during leisure, and driving), and screening for breast and/or bowel cancer (binary) as categorical variables, as well as physical activity (MET-hours), body mass index (kg/m^2^), and diet (healthy diet score, 0-7 scale) as continuous variables.

**Figure 4.**
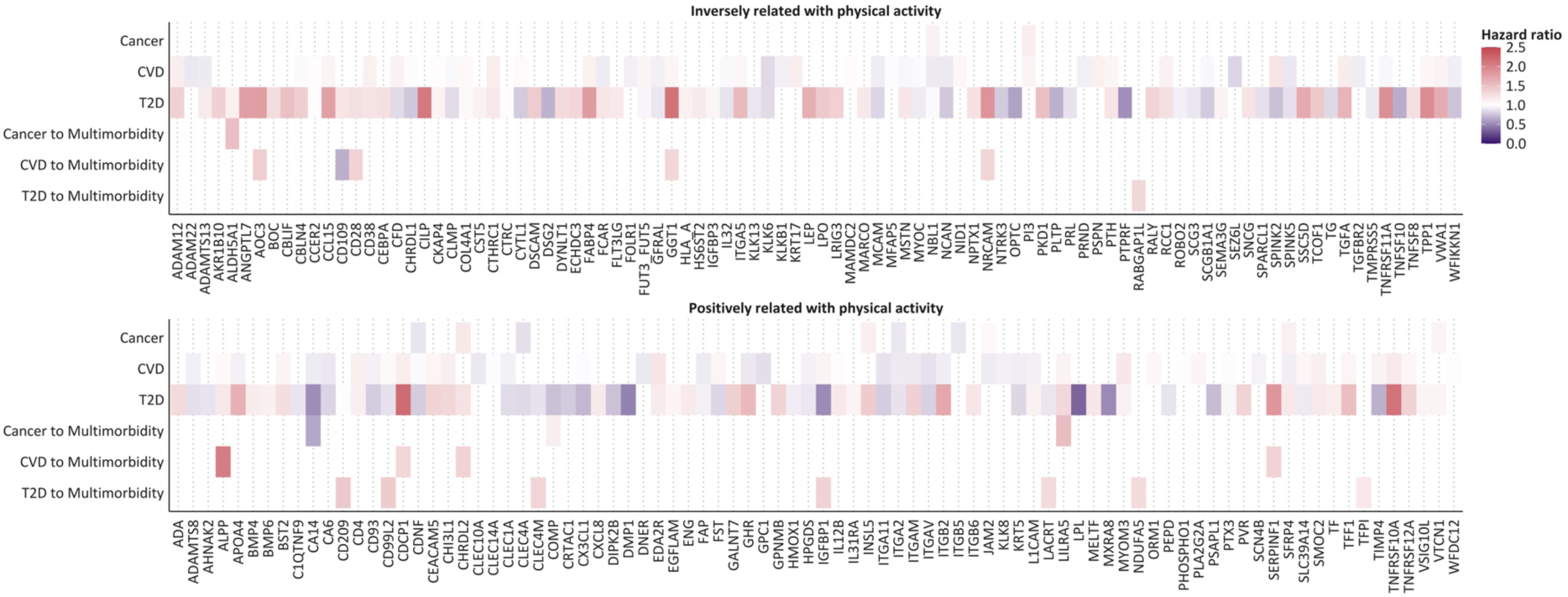
Hazard ratios for related proteins after FDR-correction. All models were stratified by age at baseline (5-year increments), sex, and country (England, Scotland, Wales), and adjusted for education level (highest, intermediate, lowest, none of those), socio-economic status (Townsend index, categorized using tertiles, missing values coded as missing), smoking (never, former, current), alcohol use (never, former, current), sedentary behavior (0-3h, 4-5h, 6-7h, >8h of daily TV watching, PC use during leisure, and driving), and screening for breast and/or bowel cancer (binary) as categorical variables, as well as physical activity (MET-hours), body mass index (kg/m^2^), and diet (healthy diet score, 0-7 scale) as continuous variables.

### Proteins related to multimorbidity

For multimorbidity following a cancer diagnosis, we identified one protein that was inversely and four proteins that were positively associated with risk of multimorbidity, while ALDH5A1 and COMP exhibited inverted U-shaped associations (both P-nonlinear<0.05). The proteomic signature was not associated with the transition from cancer to multimorbidity (HR: 1.11; 95% CI: 0.76, 1.60; q=8.10×10^−1^). For the transition from CVD to multimorbidity, eight proteins were associated with increased risk and one protein was associated with decreased risk, with AOC emerging as the top hit (HR: 1.39; 95% CI: 1.13, 1.70; q=1.29×10^−2^). We noted a tendency toward an inverse association between the proteomic signature and multimorbidity following CVD (HR: 0.76; 95% CI: 0.58, 0.99; q=1.52×10^−1^). Among individuals with T2D, eight proteins were positively associated with multimorbidity, with CD209 being the top hit (linearly associated; HR: 1.43; 95% CI: 1.16, 1.76; q=5.13×10^−3^). However, the proteomic signature was not associated with multimorbidity in this group (HR: 1.36; 95% CI: 0.88, 2.11; q=1.83×10^−1^) (Figures 3 and 4, Supplementary Figure 3).

Among the proteins inversely related to physical activity, only GGT1 was associated with single diseases (CVD, T2D) as well as multimorbidity (Figure 4, Figure 5A, Supplementary Table 3). We noted a non-linear positive association with T2D risk, and linear positive associations with CVD risk and multimorbidity risk following a CVD diagnosis (e.g., multimorbidity, HR: 1.32; 95% CI: 1.12, 1.57; q=1.50×10^−2^). Among the proteins positively associated with physical activity, we found six that were associated with single diseases (CVD, T2D) as well as multimorbidity; and one protein (CHRDL2) that was associated with cancer, T2D, and multimorbidity (Figure 4, Figure 5B, Supplementary Table 3). Across outcomes, some proteins showed consistent associations. For example, ALPP showed positive associations with both CVD and T2D (e.g., T2D, HR: 1.18; 95% CI: 1.10, 1.26; q=1.10×10^−5^) and showed even stronger associations with multimorbidity (after CVD diagnosis, HR: 2.05; 95% CI: 1.34, 3.12; q=2.50×10^−2^). However, some proteins demonstrated divergent patterns. For example, IGFBP1 was strongly inversely associated with T2D risk (HR: 0.52; 95% CI: 0.47, 0.56; q=1.20×10^−50^) but was positively associated with CVD risk (HR: 1.08; 95% CI: 1.03, 1.14; q=1.70×10^−2^) and with multimorbidity following T2D (HR: 1.37; 95% CI: 1.09, 1.73; q=3.60×10^−2^). HR curves for linear and non-linear trends are visualized in Supplementary Figure 3 for the proteomic signatures and top hits for each transition to multimorbidity.

**Figure 5.**
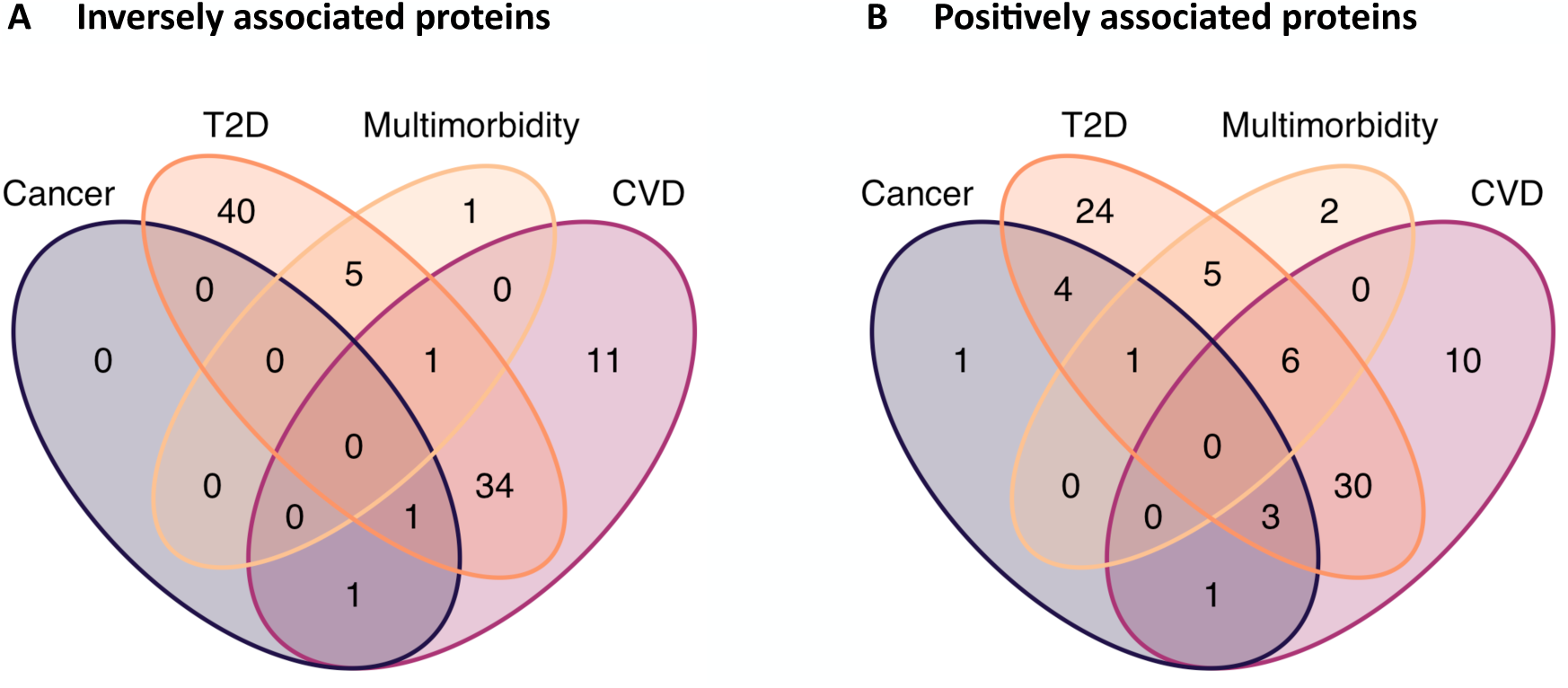
Venn diagrams for (A) inversely with physical activity associated proteins and (B) positively associated proteins for each outcome. CVD: cardiovascular disease; T2D: type 2 diabetes

### Additional analyses

We assessed the association between the proteomic signature and all outcomes after exclusion of physical activity from the models and observed minimal impact on model performance, as indicated by negligible changes in performance metrics across outcomes (Supplementary Table 4). Furthermore, mutual adjustment did not substantially alter the estimates for the proteomic signature but slightly attenuated the associations between physical activity and both T2D and multimorbidity following CVD. We saw stronger associations for the signature than for physical activity; slightly for cancer and more pronounced with non-overlapping CIs for T2D risk (Supplementary Table 5).

## 4. Discussion

In our study, we identified 223 proteins that were strongly associated with physical activity after rigorous correction for multiple testing. Second, a proteomic signature score based on these proteins, along with many individual proteins comprising the score, was associated with the risk of first incident cancer, CVD, and T2D, after adjustment for physical activity and major risk factors. Individual proteins, but not the signature score, were associated with multimorbidity risk. Third, functional enrichment analysis revealed pathways involved in maintaining tissue integrity, regulating immune and metabolic processes, and facilitating tissue repair. These mechanistic insights reinforce the role of physical activity in reducing disease burden.

Epidemiologic studies have found strong associations between physical activity and cancer and cardiometabolic diseases.^8^ The main pathways whereby physical activity contributes to better health are well-established, including improvements in insulin sensitivity, reduction of chronic low-grade inflammation, and modulation of sex hormones.^10^ In our study, we leveraged large-scale proteomic data to uncover additional, more specific molecular insights.

### Pathways and mechanisms of physical activity proteins

We found inverse associations with proteins involved in food intake, metabolism, as well as muscle and cell growth regulation, including LEP,^38^ MSTN,^39^ TGFBR2,^40^ and related proteins such as IGFBP3 and PTH. The inverse association between physical activity and pro-inflammatory LEP has been reported in other proteomic signatures of physical activity.^14,15^ A previous meta-analysis showed that physical activity may decrease LEP levels in individuals with disturbed glycemic control.^41^ Consistent with this result, we found the strongest protein association between physical activity and LEP, which is strongly associated with obesity.^42^ However, since body mass index was accounted for in the protein identification, LEP is likely related to physical activity, independently of body fat.

Conversely, physical activity was positively associated with levels of COMP, ITGAM, and MYOM3, which interact with other integrins (e.g., ITGAV, ITGB2, ITGA2, TGB5), highlighting involvement in immune cell adhesion, migration, and cartilage and muscle integrity.^43,44^ COMP^45^ and MYOM3^46^ are known to be highly responsive to physical activity.

Pathway enrichment analysis supported these results, indicating enrichment in pathways related to cell adhesion, immune function, and extracellular interactions. Proteins inversely associated with physical activity were linked to the extracellular region and multicellular processes. The enrichment of integrin complexes, receptor signaling, and opsonin binding suggests roles in immune recognition, extracellular matrix remodeling, and tissue repair. Integrins facilitate the adhesion and migration of immune cells to sites of inflammation and play a key role in suppression or promotion of tumor growth.^47^ Opsonins enhance the immune response by marking pathogens.^48^ These findings collectively suggest that physical activity may prevent disease by enhancing structural integrity of tissue and immune resilience.

### Proteomic signature and disease risk

Our analysis of physical activity-related proteins identified distinct links with the risks of cancer, CVD, T2D, and progression to multimorbidity. Higher proteomic signature scores for physical activity were associated with reduced risks of cancer, CVD, and particularly T2D, supporting the hypothesis that physical activity modulates the blood plasma proteome by regulating specific proteins that may contribute to favorable health outcomes. The strongest associations were observed for T2D, with clusters of proteins positively associated with physical activity being inversely related to T2D risk, and vice versa, contributing to a clear inverse association between the proteomic signature and T2D. Regarding multimorbidity, the proteomic signature tended to be inversely associated with the development of multimorbidity when CVD was the first detected disease but showed no association when cancer or T2D was the initial condition.

### Proteins upregulated by physical activity

We identified key proteins that may contribute to inverse overall associations with cancer, CVD, and T2D. As previously described, we found positive relations between physical activity and integrins such as ITGA2, ITGB5, and ITGA11. These three proteins were inversely associated with cancer risk, potentially indicating a yet unknown mechanism between physical activity and proteins from the integrin complex, which generally functions as context-dependent tumor suppressors.^47^ However, ITGA2 and ITGB5 are pivotal in promoting tumor cell growth and metastasis, and these proteins are often overexpressed in specific cancer tissues.^49,50^ In addition, CLEC4A showed a strong inverse association with cancer risk. CLEC4A is an inhibitory receptor that plays a crucial role in regulating both innate and adaptive immune responses, particularly in the context of inflammatory conditions.^51^ Nevertheless, there is currently no strong evidence connecting physical activity to the modulation of CLEC4A expression, but our results are supported by another recent UK Biobank study that found this protein associated with device-based and various self-reported measures of physical activity.^16^

Inverse associations with T2D risk were observed for MXRA8, LPL, IGFBP1, and DMP1. Importantly, LPL, IGFBP1, and DMP1 are directly involved in metabolic regulation, impacting lipid and glucose metabolism, and glucose homeostasis. Of these proteins, LPL demonstrates a particularly robust response to physical activity.^52,53^ On the other hand, MXRA8, also referred to as DICAM, has been shown to enhance endothelial cell adhesion and migration, potentially functioning as a negative regulator of angiogenesis,^54^ and has been linked to decreased abdominal fat.^55^

### Proteins downregulated by physical activity

We found various proteins that were inversely associated with physical activity and positively associated with disease risk. Concerning CVD, half of the top hit proteins were inversely associated with physical activity, which were, in turn, (non-linearly) positively associated with CVD risk. For example, CD38 and TGFA are involved in chronic inflammatory processes and endothelial function and play pivotal roles in blood pressure elevation and vascular dysfunction.^56,57^

Given the strong inverse correlation between physical activity and LEP, it is noteworthy that elevated LEP levels were associated with an increased risk of T2D. These findings support the notion of a physical activity-mediated pathway toward decreased T2D risk by lower circulating LEP levels.^58,59^ Moreover, GGT1 was positively associated with T2D risk, consistent with meta-analytical evidence showing that higher serum levels of GGT1 contribute to increased T2D risk.^60^ However, Mendelian randomization analyses have not substantiated the association.^61^ CILP also showed an inverse association with physical activity but was generally positively associated with T2D. Proteins from the CILP family are predominantly expressed in cartilage cells but are also found in various other tissues, and elevated serum levels have been linked to impaired glucose metabolism.^62,63^

Concerning cancer risk, PI3 (also referred to as Elafin) is typically expressed at low levels under normal physiological conditions but it is highly upregulated in response to proinflammatory cytokines.^64^ It has well-documented anti-inflammatory and antimicrobial properties through the inhibition of serine proteases involved in inflammation and bacterial defense.^65^ In addition, PI3/Elafin expression is downregulated in cancer, while its overexpression inhibits cancer cell proliferation.^66,67^ These mechanisms suggest that the observed positive association between PI3/Elafin and cancer – also reported in a previous UK Biobank study^68^ – may reflect broader inflammatory processes rather than a causal relationship between elevated PI3/Elafin and cancer development.

### Proteins associated with multimorbidity

Findings for proteins associated with multimorbidity generally exhibited greater heterogeneity. While the overall direction of association for proteins inversely related to physical activity appeared plausible, with elevated protein levels generally associated with elevated risk (particularly for GGT1 that was related to multiple outcomes), the pattern was less consistent for proteins positively associated with physical activity. Some of these proteins exhibited a decreased risk, while others exhibited an elevated risk. Some proteins demonstrated divergent associations across outcomes. For example, IGFBP1 (positively associated with physical activity) was inversely associated with T2D risk but positively with CVD and multimorbidity following T2D. Thus, IGFBP1 may be beneficial for metabolic health, as previously discussed, but potentially is adverse for cardiovascular health. This dual association has been noted in the literature, which suggests that low IGFBP1 levels indicate cardiometabolic risk, whereas higher levels are associated with worse cardiovascular outcomes.^69,70^ Lastly, certain proteins, such as ALPP, demonstrated an amplified risk for multimorbidity compared with individual disease risks, suggesting elevated risk of additional conditions in comorbid individuals.

Our results indicate that some of the identified proteins may play a role in the progression from single diseases to multimorbidity, indicating the presence of shared underlying mechanisms. However, certain pathways may exert opposing effect, emphasizing the complexity of multimorbidity as a disease state. These observations highlight the importance of a holistic approach to managing the associated disease burden.

### Proteins with divergent associations

Paradoxically, some proteins exhibited divergent associations with physical activity and disease risk. For example, half of the top hit proteins for CVD were positively associated with both physical activity and CVD risk. These included tumor necrosis factor (TNF) family members EDA2R, TNFRSF12A, and TNFRSF10A, which have also been associated with increased CVD risk in previous studies from the UK Biobank^68,71^ and the China Kadoorie Biobank.^71^ In addition to its role as an inflammatory signalling TNF receptor, EDA2R – also known as TNFRSF27 – functions as a marker of cancer-induced muscle atrophy and has been implicated in obesity-related glucose intolerance through inflammatory pathways involving c-Jun amino-terminal kinases (JNKs), which are activated via EDA2R.^72,73^ Additionally, the top hit for cancer risk was CHRDL2, which exhibited both increased cancer risk and a positive relation with physical activity. CHRDL2 is a known oncogene in gastrointestinal cancers.^74,75^

These divergent findings highlight the complexity of proteome-wide association studies and the need for cautious interpretation. While a positive relationship with physical activity may suggest a potentially protective role, elevated levels of certain proteins can also reflect underlying biological processes, such as inflammation, stress responses, or tissue damage, that may independently contribute to elevated disease risk.

### Strengths and limitations

Our study has several strengths. First, we examined the relationship between proteins, physical activity, and disease risk in an unprecedentedly large study encompassing 2,911 proteins. Second, to minimize potential reverse causation affecting protein levels, we excluded prevalent and incident cases occurring within the first year of follow-up. Third, we identified physical activity-related proteins using a rigorous approach that included comprehensive quality assessment of protein measurements, a two-step feature selection process to mitigate collider bias, consideration of non-linearity, and nested cross-validation to assess model performance. However, our study had also limitations. We relied on self-reported physical activity from a validated questionnaire, whose measurements are prone to exposure misclassification, potentially attenuating true associations between physical activity and circulating proteins. Additionally, analyzing individual proteins likely oversimplified their complex interactions, potentially underestimating their collective role in disease pathways. We addressed this concern by examining a proteomic signature as the linear combination of single proteins, as well as by conducting functional enrichment and interaction network analyses. The diverging patterns of associations between protein–physical activity and protein–outcome relationships may be the result of common limitations of observational research, prone to residual confounding and reverse causality. Lastly, the study participants are of predominantly European ancestry, limiting the generalizability of the findings to other populations.

## 5. Conclusion

We identified several proteins associated with physical activity that were also associated with cancer, CVD, T2D and the multimorbidity of these conditions. Physical activity-induced modulation of proteomic pathways involved in cell adhesion, immune surveillance, and extracellular remodelling may contribute to enhanced tissue integrity and reduced disease risk. These findings provide novel insights into the biologic mechanisms whereby physical activity may exert its beneficial effects on health.

## Declarations

### Ethics approval and consent to participate

Ethical approval was obtained from the North West Multi-Centre Research Ethics Committee. All participants provided written informed consent. This study has been conducted in line with the Declaration of Helsinki.

### Consent for publication

Not applicable.

### Availability of data and materials

UK Biobank is an open access resource. Bona fide researchers can apply to use the UK Biobank dataset by registering and applying at http://ukbiobank.ac.uk/register-apply/.

### Author Contributions

MJS and HF had full access to all the data in the study and take responsibility for the integrity of the data and the accuracy of the data analysis. Study design: MJS, VV, and HF; Acquisition, analysis, or interpretation of the data: MJS, HB, PB, RC, PF, BF, CMF, MJG, LPN, DW, MFL, VV, and HF; Manuscript writing: MJS, MFL, and HF; Critical revision of the manuscript for important intellectual content: HB, RC, PF, BF, CMF, MJG, MFL, VV, and HF.

## Acknowledgements

This research has been conducted using the UK Biobank Resource under Application Number 55870 and we express our gratitude to the participants and those involved in building the resource. This work uses data provided by patients and collected by the NHS as part of their care and support, therefore for the data linkage we want to acknowledge NHS England (Copyright © (2023), NHS England. Re-used with the permission of the NHS England [and/or UK Biobank]. All rights reserved). This research used data assets made available by National Safe Haven as part of the Data and Connectivity National Core Study, led by Health Data Research UK in partnership with the Office for National Statistics and funded by UK Research and Innovation (research which commenced between 1st October 2020–31st March 2021 grant ref MC_PC_20029; 1st April 2021–30th September 2022 grant ref MC_PC_20058).

## Disclaimer

Where authors are identified as personnel of the International Agency for Research on Cancer/ World Health Organization, the authors alone are responsible for the views expressed in this article and they do not necessarily represent the decisions, policy or views of the International Agency for Research on Cancer/ World Health Organization.

