## Supplementary material for "Proteomics signature of physical activity and risk of multimorbidity of cancer and cardiometabolic diseases"

Content

### Supplementary Table 1. ICD codes for cancer, cardiovascular diesease, and diabetes

| **Cancer** | **ICD-10 codes** | **ICD-O-3 codes** |
| --- | --- | --- |
| Esophagus (adeno) | C15 | 8140, 8141, 8143–8145, 8190–8231, 8260–8263, 8310, 8401, 8480–8490, 8550–8551, 8570–8574, 8576 |
| Stomach (cardia) | C16.0 | 8140–8145, 8147, 8210, 8211, 8214, 8220, 8221, 8230, 8231, 8255, 8260–8263, 8310, 8480, 8481, 8490, 8510, 8560, 8562, 8570–8576 |
| Colorectum | C18–C20 |  |
| Breast | C50 |  |
| Corpus uteri | C54.0, C54.1, C54.2, C54.3, C54.9, C55 |  |
| Kidney (renal cell) | C64 | 8050, 8140, 8260, 8270, 8280–8312, 8316–8320, 8340–8344 |
| Bladder | C67 |  |
| **Cardiovascular disease** | | |
| Angina pectoris | I20 |  |
| Acute myocardial infarction | I21 |  |
| Other acute ischemic heart diseases | I24 |  |
| Chronic ischemic heart diseases | I25 |  |
| Atrial fibrillation | I48 |  |
| Other cardiac arrhythmias | I49 |  |
| Heart failure: | I50 |  |
| Cerebrovascular diseases (incl. stroke) | I60 to I69 |  |
| Atherosclerosis | I70 |  |
| Other peripheral vascular diseases | I73 |  |
| **Type 2 diabetes** | E11 |  |
| ICD: international classification of diseases, SCC: squamous cell carcinoma | | |

### Supplementary Table 2. LASSO coefficients for physical activity-related proteins

| **Protein** | **Coefficient** |
| --- | --- |
| LEP | -1.49E-01 |
| MSTN | -5.24E-02 |
| TGFBR2 | -4.18E-02 |
| CHRDL1 | -3.00E-02 |
| HS6ST2 | -2.84E-02 |
| MCAM | -2.81E-02 |
| MYOC | -2.63E-02 |
| CBLN4 | -2.39E-02 |
| CD38 | -2.22E-02 |
| CLMP | -2.19E-02 |
| NBL1 | -2.12E-02 |
| AOC3 | -2.09E-02 |
| IL32 | -2.02E-02 |
| GGT1 | -1.95E-02 |
| MARCO | -1.84E-02 |
| FLT3LG | -1.72E-02 |
| SCGB1A1 | -1.46E-02 |
| PTPRF | -1.45E-02 |
| TNFRSF10C | -1.42E-02 |
| GFRAL | -1.29E-02 |
| NCAN | -1.26E-02 |
| MAMDC2 | -1.20E-02 |
| CILP | -1.18E-02 |
| DSG2 | -1.16E-02 |
| SPINK5 | -1.12E-02 |
| CST5 | -1.02E-02 |
| NTRK3 | -9.55E-03 |
| NRCAM | -9.52E-03 |
| PSPN | -9.44E-03 |
| TMPRSS5 | -8.98E-03 |
| CTRC | -8.37E-03 |
| AFP | -8.19E-03 |
| ADAMTS13 | -8.16E-03 |
| PRL | -7.96E-03 |
| TCOF1 | -7.71E-03 |
| PRND | -7.09E-03 |
| CBLIF | -6.79E-03 |
| PTH | -6.27E-03 |
| FOLR1 | -6.15E-03 |
| SPRR3 | -6.10E-03 |
| LPO | -5.92E-03 |
| KLK13 | -5.85E-03 |
| TNFSF8 | -5.79E-03 |
| CYTL1 | -5.57E-03 |
| DRAXIN | -5.49E-03 |
| SCG3 | -5.46E-03 |
| ALDH5A1 | -4.96E-03 |
| SPINK2 | -4.94E-03 |
| WFIKKN1 | -4.93E-03 |
| KRT17 | -4.69E-03 |
| VWA1 | -4.64E-03 |
| SSC5D | -4.59E-03 |
| TNFSF10 | -4.54E-03 |
| CTHRC1 | -4.51E-03 |
| TGFA | -4.46E-03 |
| CFD | -4.29E-03 |
| FUT3_FUT5 | -4.17E-03 |
| CCER2 | -3.88E-03 |
| VIT | -3.88E-03 |
| NPTX1 | -3.74E-03 |
| TG | -3.65E-03 |
| DSCAM | -3.50E-03 |
| PLTP | -3.47E-03 |
| DYNLT1 | -3.22E-03 |
| PI3 | -3.12E-03 |
| ADAM22 | -3.05E-03 |
| FABP4 | -2.74E-03 |
| CCL15 | -2.73E-03 |
| ADAM12 | -2.57E-03 |
| ECHDC3 | -2.45E-03 |
| KLK6 | -2.29E-03 |
| EXOSC10 | -2.26E-03 |
| COL28A1 | -2.19E-03 |
| PKD1 | -2.16E-03 |
| NID1 | -2.12E-03 |
| FCAR | -2.08E-03 |
| ITGA5 | -2.03E-03 |
| HLA_A | -1.97E-03 |
| RALY | -1.97E-03 |
| TNFRSF11A | -1.91E-03 |
| COL4A1 | -1.86E-03 |
| ROBO2 | -1.85E-03 |
| VWA5A | -1.80E-03 |
| ERCC1 | -1.76E-03 |
| SEZ6L | -1.71E-03 |
| NLGN2 | -1.71E-03 |
| OPTC | -1.69E-03 |
| MFAP5 | -1.69E-03 |
| RCC1 | -1.69E-03 |
| RNF4 | -1.63E-03 |
| ANXA1 | -1.59E-03 |
| RABGAP1L | -1.56E-03 |
| MMUT | -1.50E-03 |
| LRIG3 | -1.45E-03 |
| SNCG | -1.27E-03 |
| SEMA3G | -1.26E-03 |
| IGFBP3 | -1.21E-03 |
| AKR1B10 | -9.68E-04 |
| PSME2 | -9.58E-04 |
| LUZP2 | -7.44E-04 |
| BOLA1 | -7.26E-04 |
| PRSS27 | -7.23E-04 |
| ANGPTL7 | -6.22E-04 |
| SPARCL1 | -5.84E-04 |
| CKAP4 | -5.71E-04 |
| ADAMTS16 | -5.44E-04 |
| KLKB1 | -5.26E-04 |
| PBXIP1 | -5.21E-04 |
| DCC | -4.68E-04 |
| BOC | -4.27E-04 |
| TPP1 | -3.82E-04 |
| CD28 | -3.67E-04 |
| KLK15 | -5.42E-05 |
| THAP12 | -4.18E-05 |
| NEDD9 | -3.90E-05 |
| CEBPA | -3.64E-05 |
| NPHS1 | -3.33E-05 |
| SUSD4 | -1.77E-05 |
| CD109 | -6.44E-06 |
| AHNAK2 | 2.63E-05 |
| NEB | 4.14E-05 |
| CHI3L1 | 1.63E-04 |
| KLK8 | 1.76E-04 |
| CXCL8 | 2.06E-04 |
| VTCN1 | 2.60E-04 |
| CLEC10A | 4.63E-04 |
| ITGB6 | 5.34E-04 |
| ALPP | 5.44E-04 |
| BST2 | 6.16E-04 |
| ORM1 | 7.34E-04 |
| TYRP1 | 9.87E-04 |
| ITM2A | 1.07E-03 |
| TIMP4 | 1.12E-03 |
| TFPI | 1.25E-03 |
| ANXA5 | 1.31E-03 |
| GPNMB | 1.38E-03 |
| BMP6 | 1.41E-03 |
| MXRA8 | 1.47E-03 |
| IL12B | 1.49E-03 |
| PHOSPHO1 | 1.51E-03 |
| SLC39A14 | 1.79E-03 |
| KIR2DL3 | 2.00E-03 |
| SFRP4 | 2.11E-03 |
| DIPK2B | 2.13E-03 |
| PSAPL1 | 2.26E-03 |
| EXTL1 | 2.34E-03 |
| FLT1 | 2.41E-03 |
| LILRA5 | 2.47E-03 |
| ITGA11 | 2.65E-03 |
| PVR | 2.73E-03 |
| NDUFA5 | 2.74E-03 |
| GHR | 2.86E-03 |
| ITGB5 | 3.00E-03 |
| DUSP13 | 3.02E-03 |
| PTX3 | 3.16E-03 |
| GP1BB | 3.17E-03 |
| CD4 | 3.33E-03 |
| ITGA2 | 3.45E-03 |
| EGFLAM | 3.63E-03 |
| SERPINF1 | 3.71E-03 |
| DOCK9 | 4.08E-03 |
| ENG | 4.21E-03 |
| CEACAM5 | 4.41E-03 |
| GALNT7 | 4.45E-03 |
| LACRT | 4.81E-03 |
| CD99L2 | 4.84E-03 |
| VSIG10L | 5.03E-03 |
| KRT5 | 5.07E-03 |
| SCN4B | 5.11E-03 |
| TNFRSF10A | 5.34E-03 |
| MELTF | 5.47E-03 |
| CDNF | 5.89E-03 |
| HPGDS | 5.92E-03 |
| ADA | 6.29E-03 |
| TF | 6.41E-03 |
| CX3CL1 | 6.71E-03 |
| GPC1 | 6.79E-03 |
| INSL5 | 6.80E-03 |
| FAP | 6.86E-03 |
| JAM2 | 7.13E-03 |
| ITGAV | 7.41E-03 |
| CDH5 | 7.45E-03 |
| HMOX1 | 7.54E-03 |
| BMP4 | 8.21E-03 |
| PODXL | 8.25E-03 |
| CD209 | 8.29E-03 |
| PEPD | 1.04E-02 |
| CD93 | 1.04E-02 |
| PLA2G2A | 1.05E-02 |
| CHRDL2 | 1.10E-02 |
| SMOC2 | 1.10E-02 |
| IL31RA | 1.12E-02 |
| NTF3 | 1.18E-02 |
| EDA2R | 1.20E-02 |
| FST | 1.24E-02 |
| ITGB2 | 1.24E-02 |
| C1QTNF9 | 1.28E-02 |
| IGFBP1 | 1.33E-02 |
| CLEC4A | 1.36E-02 |
| SPINK6 | 1.36E-02 |
| WFDC12 | 1.43E-02 |
| CA6 | 1.46E-02 |
| CLEC4M | 1.54E-02 |
| DNER | 1.64E-02 |
| CDCP1 | 1.78E-02 |
| CRTAC1 | 1.87E-02 |
| DMP1 | 1.94E-02 |
| TFF1 | 2.01E-02 |
| ADAMTS8 | 2.07E-02 |
| BAG3 | 2.13E-02 |
| CXCL14 | 2.15E-02 |
| MEGF10 | 2.18E-02 |
| ADAMTSL5 | 2.34E-02 |
| TNFRSF12A | 2.35E-02 |
| APOA4 | 2.44E-02 |
| CLEC1A | 2.51E-02 |
| L1CAM | 2.77E-02 |
| CLEC14A | 2.97E-02 |
| CA14 | 3.47E-02 |
| LPL | 3.81E-02 |
| MYOM3 | 3.88E-02 |
| ITGAM | 4.19E-02 |
| COMP | 5.73E-02 |

### Supplementary Table 3. Proteins associated with diseases as well as multimorbiditiy

| **Protein** | **Transition** | **HR (95% CI)** | **q** | **P-nonlinear** |
| --- | --- | --- | --- | --- |
| **Inversely associated with physical activity** | | |  |  |
| GGT1 | Baseline to T2D | 2.14 (1.84, 2.49) | 7.80E-51 | 4.80E-05 |
|  | Baseline to CVD | 1.07 (1.03, 1.12) | 1.60E-02 | 4.30E-01 |
|  | CVD to Multimorbidity | 1.32 (1.12, 1.57) | 1.50E-02 | 6.40E-01 |
| **Positively associated with physical activity** | | |  |  |
| ALPP | Baseline to CVD | 1.04 (0.96, 1.13) | 9.30E-05 | 1.10E-04 |
|  | Baseline to T2D | 1.18 (1.10, 1.26) | 1.10E-05 | 5.30E-01 |
|  | CVD to Multimorbidity | 2.05 (1.34, 3.12) | 2.50E-02 | 4.10E-03 |
| CA14 | Baseline to CVD | 0.92 (0.84, 1.01) | 3.10E-04 | 1.30E-02 |
|  | Baseline to T2D | 0.53 (0.45, 0.61) | 4.90E-32 | 7.30E-04 |
|  | Cancer to Multimorbidity | 0.64 (0.47, 0.87) | 2.20E-02 | 5.70E-01 |
| CD99L2 | Baseline to CVD | 1.05 (0.96, 1.14) | 6.60E-03 | 1.40E-02 |
|  | Baseline to T2D | 0.88 (0.82, 0.94) | 2.30E-03 | 8.50E-02 |
|  | T2D to Multimorbidity | 1.42 (1.14, 1.76) | 1.00E-02 | 3.20E-01 |
| CDCP1 | Baseline to CVD | 1.15 (1.10, 1.20) | 8.80E-08 | 9.50E-02 |
|  | Baseline to T2D | 2.19 (1.88, 2.55) | 1.20E-50 | 5.50E-06 |
|  | CVD to Multimorbidity | 1.32 (1.09, 1.60) | 2.60E-02 | 1.40E-01 |
| CHRDL2 | Baseline to Cancer | 1.18 (1.10, 1.27) | 8.20E-05 | 7.60E-01 |
|  | Baseline to T2D | 1.22 (1.14, 1.31) | 4.10E-07 | 6.30E-01 |
|  | CVD to Multimorbidity | 1.35 (1.10, 1.67) | 2.60E-02 | 2.80E-01 |
| IGFBP1 | Baseline to CVD | 1.08 (1.03, 1.14) | 1.70E-02 | 6.40E-01 |
|  | Baseline to T2D | 0.52 (0.47, 0.56) | 1.20E-50 | 4.10E-01 |
|  | T2D to Multimorbidity | 1.37 (1.09, 1.73) | 3.60E-02 | 3.90E-01 |
| LILRA5 | Baseline to CVD | 1.10 (1.05, 1.15) | 5.00E-04 | 2.00E-01 |
|  | Baseline to T2D | 1.31 (1.22, 1.41) | 1.70E-11 | 1.40E-01 |
|  | Cancer to Multimorbidity | 1.53 (1.11, 2.13) | 4.90E-02 | 9.50E-01 |
|  | Baseline to CVD | 1.04 (0.96, 1.13) | 9.30E-05 | 1.10E-04 |
| CI: confidence interval; CVD: cardiovascular disease; HR: hazard ratio; T2D: type 2 diabetes  All models were stratified by age at baseline (5-year increments), sex, and country (England, Scotland, Wales), and adjusted for education level (highest, intermediate, lowest, none of those), socio-economic status (Townsend index, categorized using tertiles, missing values coded as missing), smoking (never, former, current), alcohol use (never, former, current), sedentary behavior (0-3h, 4-5h, 6-7h, >8h of daily TV watching, PC use during leisure, and driving), and screening for breast and/or bowel cancer (binary) as categorical variables, as well as physical activity (MET-hours), body mass index (kg/m2), and diet (healthy diet score, 0-7 scale) as continuous variables. | | | | |

### Supplementary Table 4. Model performance metrics for single and mutually adjusted models for the proteomics signature and physical activity

| **Outcome** | **Base model** | **Model with signature** | **Model with MVPA** | **Model with signature + MVPA** |
| --- | --- | --- | --- | --- |
| **Baseline to cancer** |  |  |  |  |
| AIC | 16007.5 | 15996.5 | 15998.6 | 15993.2 |
| LRT X^2^ | 44.4 | 57.3 | 55.2 | 62.6 |
| Overall P-value | 6.92×10^-3^ | 2.45×10^-4^ | 4.62×10^-4^ | 7.47×10^-5^ |
| C-index | 0.715 | 0.712 | 0.714 | 0.713 |
| **Baseline to CVD** |  |  |  |  |
| AIC | 49489.9 | 49490.3 | 49491.4 | 49492.2 |
| LRT X^2^ | 264.3 | 265.9 | 264.8 | 266.0 |
| Overall P-value | <1.00×10^-30^ | <1.00×10^-30^ | <1.00×10^-30^ | <1.00×10^-30^ |
| C-index | 0.658 | 0.658 | 0.658 | 0.658 |
| **Baseline to T2D** |  |  |  |  |
| AIC | 18695.4 | 18602.4 | 18686.2 | 18604.3 |
| LRT X^2^ | 1238.5 | 1333.4 | 1249.7 | 1333.5 |
| Overall P-value | <1.00×10^-30^ | <1.00×10^-30^ | <1.00×10^-30^ | <1.00×10^-30^ |
| C-index | 0.805 | 0.812 | 0.805 | 0.812 |
| **Cancer to multimorbidity** |  |  |  |  |
| AIC | 680.1 | 682.0 | 682.0 | 683.8 |
| LRT X^2^ | 23.3 | 23.5 | 23.4 | 23.7 |
| Overall P-value | 4.42×10^-1^ | 4.91×10^-1^ | 4.95×10^-1^ | 5.36×10^-1^ |
| C-index | 0.760 | 0.760 | 0.760 | 0.762 |
| **CVD to multimorbidity** |  |  |  |  |
| AIC | 1429.0 | 1426.3 | 1430.3 | 1428.3 |
| LRT X^2^ | 74.1 | 78.8 | 74.8 | 78.8 |
| Overall P-value | 2.81×10^-7^ | 9.53×10^-8^ | 4.03×10^-7^ | 1.76×10^-7^ |
| C-index | 0.795 | 0.796 | 0.797 | 0.796 |
| **T2D to multimorbidity** |  |  |  |  |
| AIC | 1287.7 | 1286.4 | 1289.7 | 1288.3 |
| LRT X^2^ | 19.4 | 26.8 | 19.4 | 26.8 |
| Overall P-value | 6.77×10^-1^ | 4.22×10^-1^ | 7.30×10^-1^ | 4.75×10^-1^ |
| C-index | 0.675 | 0.689 | 0.675 | 0.689 |
| AIC: Akaike information criterion; CVD: cardiovascular disease; LRT: log-likelihood ratio test; MVPA: moderate-to-vigorous physical activity; T2D: type 2 diabetes.  All models were stratified by age at baseline (5-year increments), sex, and country (England, Scotland, Wales), and adjusted for education level (highest, intermediate, lowest, none of those), socio-economic status (Townsend index, categorized using tertiles, missing values coded as missing), smoking (never, former, current), alcohol use (never, former, current), sedentary behavior (0-3h, 4-5h, 6-7h, >8h of daily TV watching, PC use during leisure, and driving), and screening for breast and/or bowel cancer (binary) as categorical variables, as well as body mass index (kg/m2), and diet (healthy diet score, 0-7 scale) as continuous variables. | | | | |

### Supplementary Table 5. Hazard ratios and 95% confidence intervals for the proteomic signature and physical activity under single and mutual adjustment scenarios

| **Outcome** | **Model adjustment** | **Signature** | **MVPA** |
| --- | --- | --- | --- |
|  |  | Hazard ratio (95% CI) | Hazard ratio (95% CI) |
| Baseline to cancer | Single | 0.84 (0.76, 0.92) | 0.90 (0.84, 0.96) |
|  | Mutual | 0.87 (0.78, 0.96) | 0.92 (0.86, 0.99) |
| Baseline to CVD | Single | 0.97 (0.92, 1.02) | 0.99 (0.96, 1.02) |
|  | Mutual | 0.97 (0.92, 1.02) | 0.99 (0.96, 1.03) |
| Baseline to T2D | Single | 0.66 (0.60, 0.72) | 0.91 (0.86, 0.96) |
|  | Mutual | 0.66 (0.60, 0.72) | 0.99 (0.94, 1.05) |
| Cancer to multimorbidity | Single | 1.08 (0.76, 1.54) | 0.97 (0.80, 1.17) |
|  | Mutual | 1.11 (0.76, 1.60) | 0.95 (0.78, 1.16) |
| CVD to multimorbidity | Single | 0.76 (0.59, 0.97) | 0.93 (0.78, 1.11) |
|  | Mutual | 0.76 (0.58, 0.99) | 0.99 (0.82, 1.18) |
| T2D to multimorbidity | Single | 1.35 (0.88, 2.08) | 1.00 (0.85, 1.17) |
|  | Mutual | 1.36 (0.88, 2.11) | 0.99 (0.84, 1.16) |
| CI: confidence interval; CVD: cardiovascular disease; MVPA: moderate-to-vigorous physical activity; T2D: type 2 diabetes.  All models were stratified by age at baseline (5-year increments), sex, and country (England, Scotland, Wales), and adjusted for education level (highest, intermediate, lowest, none of those), socio-economic status (Townsend index, categorized using tertiles, missing values coded as missing), smoking (never, former, current), alcohol use (never, former, current), sedentary behavior (0-3h, 4-5h, 6-7h, >8h of daily TV watching, PC use during leisure, and driving), and screening for breast and/or bowel cancer (binary) as categorical variables, as well as body mass index (kg/m^2^), and diet (healthy diet score, 0-7 scale) as continuous variables. | | | |

#### Supplementary Figure 1. Flowchart of participant inclusion

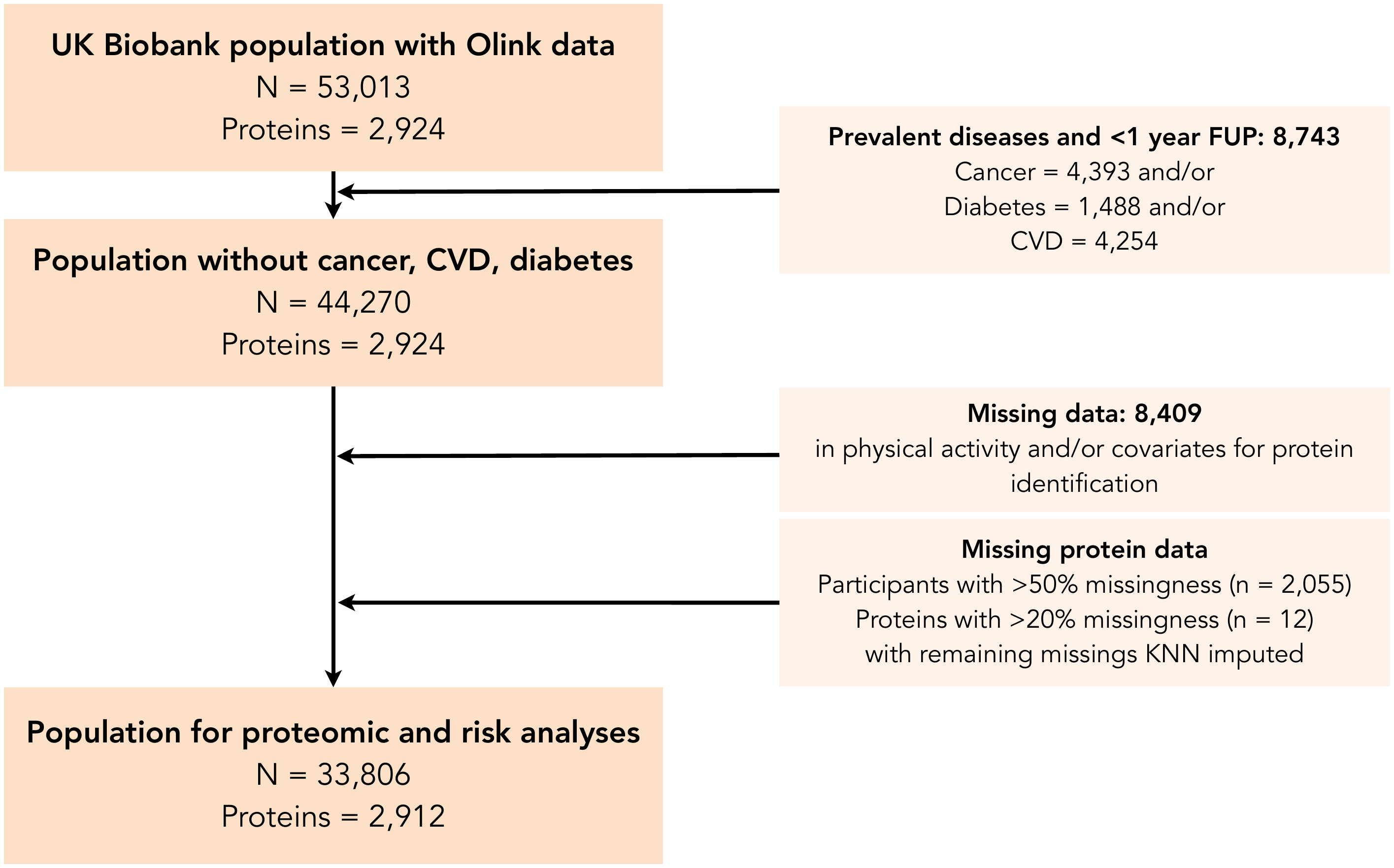

#### Supplementary Figure 2. Continuous hazard ratios for the top hits among all proteins for the baseline–disease transitions

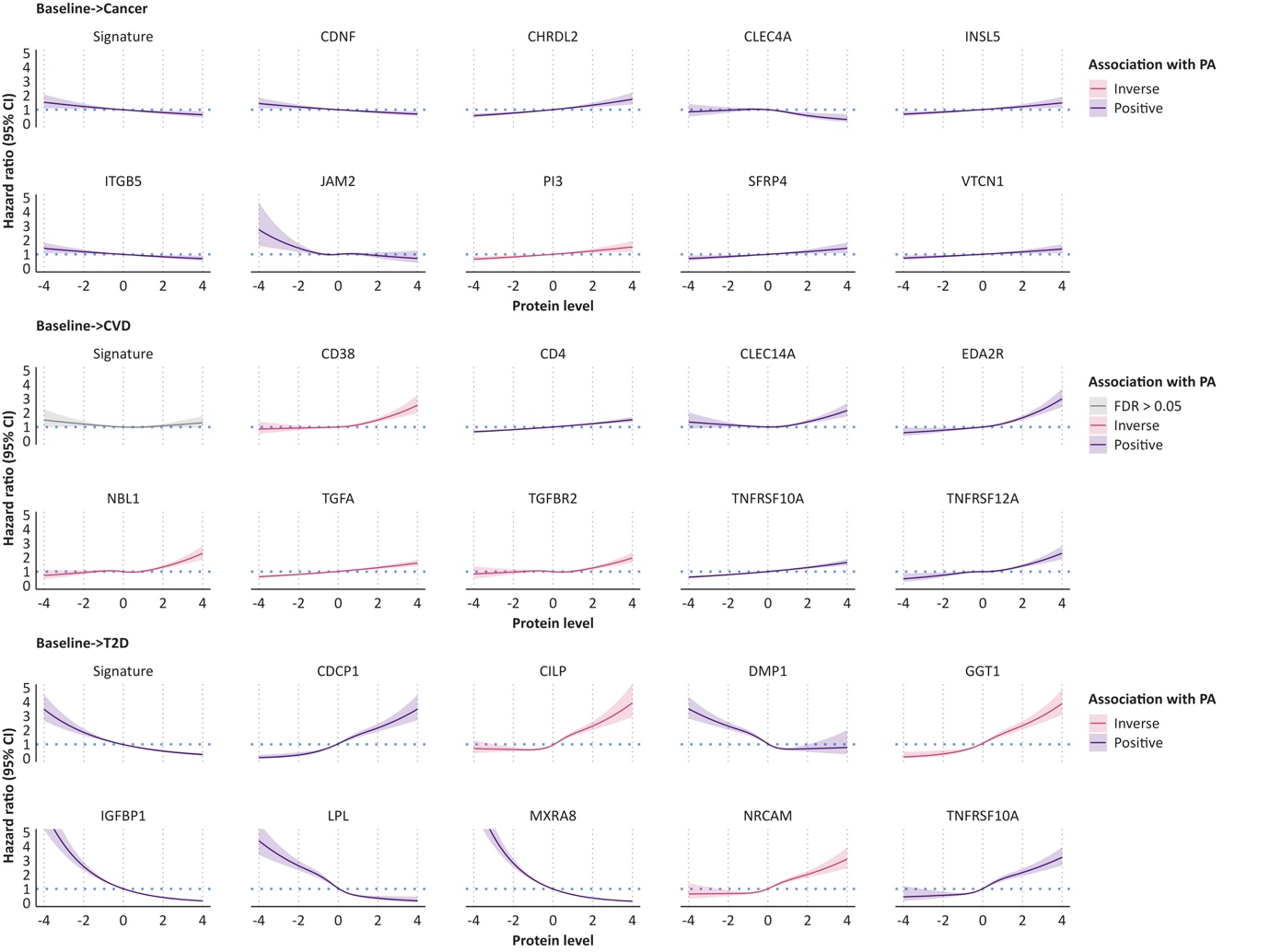

#### Supplementary Figure 3. Continuous hazard ratios for the top hits among all proteins for the disease–multimorbidity transitions

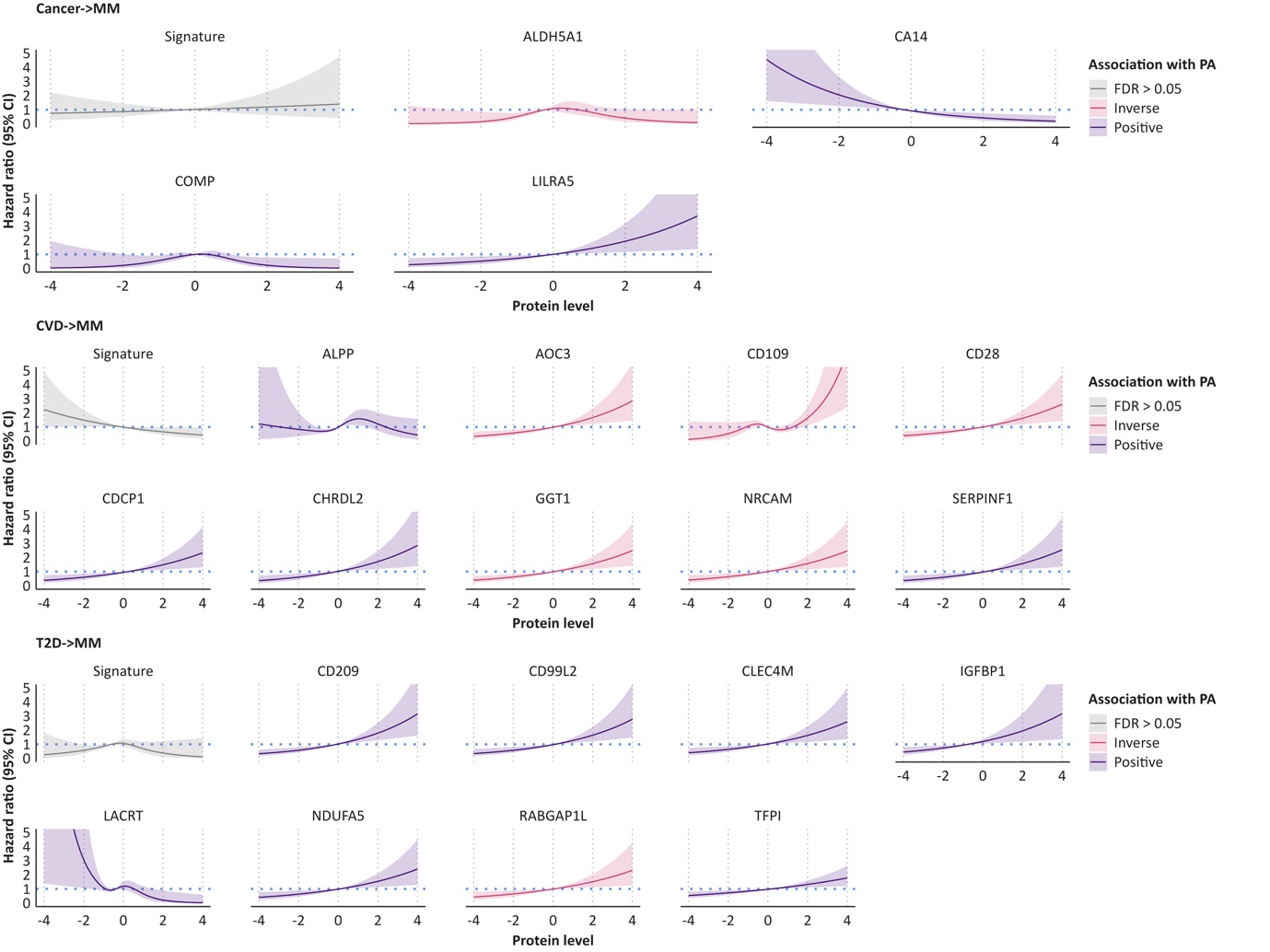
